# Isolated great saphenous vein stripping for the treatment of varicose veins in lower limbs: a prospective study

**DOI:** 10.64898/2026.06.17.26355878

**Authors:** Felipe Soares Oliveira Portela, Andressa Cristina Sposato Louzada, Maria Fernanda Cassino Portugal, Marcelo Fiorelli Alexandrino da Silva, Lucas Lembrança, Bruno Fabricio Feio Antunes, Alexandre Fioranelli, Nelson Wolosker

**Author notes:** **Corresponding author:** Felipe Soares Oliveira Portela, Department of Vascular Surgery, Hospital Israelita Albert Einstein - Avenida Albert Einstein, 627, bloco A1, sala 423, Morumbi. ZIP:05652-900, São Paulo, SP – Brazil., **, **. The authors have no conflicts of interest to declare. **Data Availability Statement:** The data that support the findings of this study are available from the corresponding author upon reasonable request.

## Abstract

**Background:** Endovenous techniques are considered the gold standard for treating great saphenous vein (GSV) insufficiency, but access remains limited in low- and middle-income countries. In such contexts, simplified conventional surgeries may represent viable alternatives. This study aimed to compare outcomes of isolated GSV stripping with conventional surgery (stripping plus varicose vein resection) in patients with varicose veins (VV) associated with GSV insufficiency.

**Methods:** A prospective interventional study was conducted including 34 patients with VV (CEAP C2–C6), divided into two groups: Conventional (C, n=17) and Isolated Saphenectomy (IS, n=17). Quality of life was assessed preoperatively and at 2 and 6 months postoperatively using the Venous Clinical Severity Score (VCSS) and VEINES-QoL/Sym questionnaires. Varicose vein evolution in the IS group was quantified using a standardized visual scoring system. Statistical analyses included Student’s t-test, chi-square, and generalized estimating equations (p≤0.05).

**Results:** Both groups were demographically comparable. Surgical treatment significantly improved VCSS and VEINES scores in both groups (p<0.005), with no intergroup difference at 6 months. In the IS group, the mean reduction in visible VV was 46% (range 20–90%). CEAP classification improved in both groups, with migration toward less severe categories postoperatively. No major complications were reported.

**Conclusion:** Isolated GSV stripping yields comparable short- and mid-term improvements in symptoms and quality of life to conventional surgery, while reducing operative extent. In resource-limited settings, this abbreviated technique may expand access to treatment for VV, improving patient outcomes and reducing healthcare costs without compromising clinical efficacy.

## INTRODUCTION

Lower limb varicose veins (VV) are the most common vascular disease worldwide, leading to significant morbidity and imposing high costs on healthcare systems.^1^ In the United States alone, it is estimated that approximately 30 million individuals are affected by VV, with annual treatment costs reaching up to US$1 billion.^2^ Besides causing symptoms such as pain, edema, and heavy sensation in the legs, untreated VV can progress to significant complications, including leg ulcers and an increased risk of deep vein thrombosis.^3–5^

Currently, endovenous thermal ablation (EVTA) techniques are considered the gold standard for treating great saphenous vein (GSV) insufficiency in patients with VV.^6,7^ However, there are significant disparities in access to these technologies, especially in middle- and low-income countries.^8^ In Brazil, for example, approximately 75% of the population relies solely on the public health system^9^, where conventional surgery still prevails as the main technique. It is estimated that about 70,000 conventional VV surgeries are performed annually in the public health system of the largest country in Latin America^10^

In cases of VV associated with GSV insufficiency, several studies have demonstrated that isolated GSV ablation, without simultaneous resection of VV, can offer good outcomes in terms of patient satisfaction and reducing the need for additional interventions^11–13^. However, there is currently no research evaluating whether similar benefits are achieved through conventional surgery involving isolated GSV resection as a treatment approach.

In low- and middle-income countries, access to treatment for VV often occurs at late stages, with patients in more advanced ages and experiencing more severe degrees of venous disease. In such contexts, abbreviated surgery may represent a viable alternative that can improve quality of life, reduce the rates of complications associated with venous disease, and not significantly increase risks for these patients.

The objective of this study was to evaluate, in patients with VV associated with GSV insufficiency, whether treatment with saphenectomy alone is comparable to conventional treatment (saphenectomy combined with varicose vein resection) in terms of quality of life, patient satisfaction, and the reduction of visible VV volume.

## METHODS

### Study Design and Ethical Issues

A prospective, interventional study was conducted involving patients with VV related to GSV insufficiency who were indicated for surgical treatment. Participants were divided into two groups:

- Conventional Group (C): Patients underwent conventional surgical treatment, which included GSV stripping along with the resection of all VV.
- Isolated Saphenectomy Group (IS): Patients underwent isolated GSV stripping without resection of visible varicose veins.

The study was approved by the Institutional Review Board (CAAE 69606917.7.3002.0083), and all participants provided informed consent. This study is registered at ClinicalTrials.gov (NCT07616115).

### Sample size calculation

Considering an average satisfaction score of 7 points (on a scale ranging from 0 to 10) for conventional treatment, and assuming a minimum expected score of 5 for the alternative procedure, with an estimated standard deviation of 2 points, statistical power of 80%, and a significance level of 95%, we estimated a minimum need for 16 participants in each group.

### Recruitment of participants and allocation to groups

Participants were recruited from a single public health institution in São Paulo, Brazil.

Inclusion criteria: individuals aged between 18 and 70, classified with VV as CEAP C2 to C6, who formally consented to participate in the study.

Exclusion criteria: individuals with small saphenous vein insufficiency; previous deep vein thrombosis; deep venous insufficiency; previous treatment of varicose veins in the target limb; or comorbidities that could affect clinical or quality of life assessment after surgery, such as orthopedic diseases (osteoarthritis, herniated disc, among others).

According to institutional guidelines, the treatment of lower limb VV is performed in stages (one limb per approach). To ensure that all patients received the recommended conventional treatment by the end of the study, patients were divided into two groups based on their conditions:

- Patients with VV and unilateral GSV insufficiency were included in the Conventional group, undergoing saphenectomy combined with the resection of VV in the affected limb in a single procedure.
- Patients with VV and bilateral GSV insufficiency were included in the Isolated Saphenectomy group. In these cases, unilateral saphenectomy was initially performed. After the study was completed, treatment was finalized with contralateral saphenectomy and removal of VV in both limbs.

### Surgical procedure

Patients were treated in the operating room under spinal anaesthesia. The treatment of the GSV was performed through an incision at the top of the thigh, which included the ligation of tributary veins and resection of the insufficient segment using a pin-stripper. The VV were resected through staggered incisions.

### Quality of life assessment (PROMs)

The assessment was performed at three different times: preoperatively, and at 2 and 6 months after surgery. Two validated instruments were utilized:

1) Venous Clinical Severity Score (VCSS)^14^ Portuguese version^15^:

The VCSS consists of 10 attributes: pain, varicose veins, edema, pigmentation, inflammation, induration, number of ulcers, duration of ulcers, size of ulcers, and compressive therapy. Each attribute is rated on a scale from 0 to 3, indicating the severity of symptoms as absent, mild, moderate, or severe. In this context, the higher score indicates more severe venous disease. All these attributes were analyzed in this study.

2) VEINES/QOL-Sym^16^

This is a specific quality-of-life assessment tool for venous disease, comprising 26 items that cover various aspects, including 10 questions about symptoms, 9 questions regarding limitations in daily activities, 5 questions on psychosocial impact, 1 item related to changes in complaints over the past year, and a question about the time of day when symptoms are most frequent. Here, a higher score signifies a better quality of life for the patient. To enable a more comprehensive analysis of changes in quality of life before and after treatment, the VEINES scores were converted into a 0-100 scale using a method previously described in the literature.^17^

### Visual documentation

It was performed at two different times: preoperatively and two months post-procedure. To document each patient’s VV, we developed a standardized form for systematic recording.

Preoperatively, we adopted a standardized marking system to better characterize the VV: small-caliber veins (<3 mm) were identified in blue, medium-caliber veins (3-5 mm) in red, and large-caliber veins (>5 mm) in black. These markings for each patient were transferred to the forms for subsequent storage and analysis.

To enable objective assessment and facilitate comparisons of the evolution of VV after isolated saphenectomy, we created a scale based on the caliber and extent of each varicose vein, ranging from 1 to 6 points according to Table 1:

**Table 1.** Standardization of scores for each varicose vein, according to the classification proposed by the authors.

| Category | Caliber | Extent | Score |
| --- | --- | --- | --- |
| I | < 3 mm | < 5 cm | 1 |
| II | < 3 mm | ≥ 5 cm | 2 |
| III | 3-5 mm | < 5 cm | 3 |
| IV | 3-5 mm | ≥ 5 cm | 4 |
| V | > 5 mm | < 5 cm | 5 |
| VI | > 5 mm | ≥ 5 cm | 6 |

Each operated limb received a final score resulting from the aggregate of the scores assigned to each vein marked on that limb. We compared the pre-operative and post-operative scores to calculate the percentage reduction in VV following isolated saphenectomy.

Additionally, we analyzed demographic data, CEAP characteristics, VCSS, VEINES-QoL, and VEINES-SYM scores both before treatment, and at 2 and 6 months after treatment. We also analyzed the quantitative changes in collateral veins.

### Statistical analysis

All statistical analyses were conducted using SPSS software version 22 (IBM, Armonk, New York, USA). The Student’s t-test was used to assess continuous variables, while the chi-square test was used to analyze categorical variables.

To evaluate variations in quality-of-life scores at each time point (both pre- and post-treatment), a generalized estimating equation with a normal distribution and an identity link function was applied. This analysis assumed an AR(1) correlation matrix between the time points and included Bonferroni corrections for multiple comparisons. Multiple linear regression was used for the multivariate analysis.

A P-value of less than or equal to 0.05 was considered statistically significant for all tests.

## RESULTS

### Demographics

A total of thirty-four patients participated in the study, with 17 assigned to each group. The demographic and clinical characteristics of the participants are shown in Table 2. The two groups were similar regarding age (early 50s), body mass index (both overweight), gender distribution (predominantly female), and prevalence of comorbidities. There were no statistically significant differences between the groups for all variables.

**Table 2.** Clinical and demographic characteristics of patients with lower limb varicose veins and great saphenous vein insufficiency undergoing surgical treatment.

| Variable | C - n | C - % | IS - n | IS - % | Total - n | Total - % | p-value |
| --- | --- | --- | --- | --- | --- | --- | --- |
| Mean age (years) | 49,24 ± 10,35 |  | 52,59 ± 7,72 |  | 50,92 ± 9,15 |  | 0,393<sup>1</sup> |
| BMI (kg/m<sup>2</sup>) | 28,74 ± 3,16 |  | 28,98 ± 4,76 |  | 28,86 ± 3,98 |  | 0,865<sup>1</sup> |
| Gender |  |  |  |  |  |  | 0,151<sup>2</sup> |
| Male | 8 | 47,1 | 4 | 23,5 | 12 | 35,3 |  |
| Female | 9 | 52,9 | 13 | 76,5 | 22 | 64,7 |  |
| Hypertension | 3 | 17,6 | 8 | 47,1 | 11 | 32,4 | 0,067<sup>2</sup> |
| Diabetes | 1 | 5,9 | 1 | 5,9 | 2 | 5,9 | >0,999<sup>3</sup> |
| Smoking |  |  |  |  |  |  | 0,164<sup>□</sup> |
| Previous | 2 | 11,8 | 1 | 5,9 | 3 | 8,8 |  |
| Current | 1 | 5,9 | 5 | 29,4 | 6 | 17,6 |  |
| Total | 17 | 100,0 | 17 | 100,0 | 34 | 100,0 |  |

## CEAP (C)

Table 3 shows the CEAP clinical classification data for patients in both groups, both before and after surgical treatment. Preoperatively, most cases were classified as CEAP C3 and C2. After surgery, there was a general clinical improvement, with a noticeable toward less severe classes (especially C1). The IS group had a relatively higher proportion in C2 in the postoperative period compared to the Conventional group. There were no significant differences in the distribution of CEAP classes between the groups.

**Table 3.** Comparison of pre- and post-operative CEAPs for the Conventional and Isolated Saphenectomy groups.

|  | Group |  |  |  |  |  |  |
| --- | --- | --- | --- | --- | --- | --- | --- |
|  | C |  | IS |  | Total |  | p |
|  | n | % | n | % | n | % |  |
| **Pre-operative CEAP** |  |  |  |  |  |  | 0,186£ |
| C2 | 5 | 29,4 | 5 | 29,4 | 10 | 29,4 |  |
| C3 | 5 | 29,4 | 9 | 52,9 | 14 | 41,2 |  |
| C4 | 5 | 29,4 | 3 | 17,6 | 8 | 23,5 |  |
| C6 | 2 | 11,8 | 0 | 0,0 | 2 | 5,9 |  |
| **Post-operative CEAP** |  |  |  |  |  |  | 0,317£ |
| C1 | 10 | 58,8 | 9 | 52,9 | 19 | 55,9 |  |
| C2 | 3 | 17,6 | 7 | 41,2 | 10 | 29,4 |  |
| C4 | 2 | 11,8 | 1 | 5,9 | 3 | 8,8 |  |
| C5 | 2 | 11,8 | 0 | 0,0 | 2 | 5,9 |  |
| £ Chi-square trend test |  |  |  |  |  |  |  |

Caption: C – Conventional group IS – Isolated Saphenectomy group

### Quality of life

Table 4 presents the quality of life scores for both groups before surgery. The VCSS was similar in both groups. However, regarding the VEINES scores, both in the quality of life and symptoms domains, patients in the Conventional group had significantly higher values (indicating better clinical condition) compared to those in the IS group.

**Table 4.** Quality of life scores for the Conventional (C) and Isolated Saphenectomy (IS) groups preoperatively.

| AVERAGE PRE-TREATMENT VALUES |  |  |  |
| --- | --- | --- | --- |
|  | **C** | **IS** | **p** |
| **VCSS** | 10,00 ± 3,57 | 8,29 ± 1,53 | 0,433 |
| **VEINES-QoL** | 69,0 ± 11,8 | 62,5 ± 14,1 | **0,021** |
| **VEINES-SYM** | 61,1 ± 13,0 | 56,1 ± 15,7 | **0,023** |

Table 5 presents the mean postoperative values of the VCSS, VEINES-QoL (quality of life), and VEINES-Sym (symptoms) scores in both groups, assessed at 2 and 6 months after treatment. Surgical treatment resulted in a significant improvement in quality of life in both groups (p < 0.005). This improvement, observed as early as 2 months postoperatively, was maintained throughout the follow-up period, lasting until the end of the 6-month follow-up. Although the IS group started with a lower quality of life, it achieved results similar to those of the group undergoing conventional treatment.

**Table 5.** Postoperative evolution of quality of life scores.

| AVERAGE POST-TREATMENT VALUES |  |  |  |
| --- | --- | --- | --- |
|  | **C** | **SI** | **p** |
| 2 months |  |  |  |
| **VCSS** | 3,12 ± 1,36 | 3,29 ± 2,29 | >0,999 |
| **VEINES-QoL** | 93,2 ± 4,2 | 90,5 ± 5,5 | >0,999 |
| **VEINES-SYM** | 93,4 ± 5,1 | 88,9 ± 8,4 | >0,999 |
| 6 months |  |  |  |
| **VCSS** | 3,00 ± 1,54 | 3,18 ± 1,85 | >0,999 |
| **VEINES-QoL** | 94,0 ± 2,5 | 91,1 ± 4,4 | 0,991 |
| **VEINES-SYM** | 93,3 ± 3,0 | 88,6 ± 6,1 | 0,557 |

### Evaluation of VV in the postoperative period of isolated GSV treatment

Table 6 shows the individual evolution of varicose veins in the 17 patients who underwent isolated saphenectomy, showing the score before and after the procedure, as well as the percentage reduction for each case. There was an average reduction of 46% in the caliber and extension of varicose veins, ranging from 20% to 90% depending on the patient.

**Table 6.** Evolution of varicose veins after treatment of the great saphenous vein alone.

| Patient | Pre-operative score | Post-operative score | Reduction (%) |
| --- | --- | --- | --- |
| 1 | 31 | 12 | -61,29 |
| 2 | 32 | 10 | -68,75 |
| 3 | 21 | 10 | -52,38 |
| 4 | 20 | 2 | -90,00 |
| 5 | 24 | 16 | -33,33 |
| 6 | 16 | 9 | -43,75 |
| 7 | 60 | 49 | -18,33 |
| 8 | 41 | 31 | -24,39 |
| 9 | 38 | 16 | -57,89 |
| 10 | 21 | 17 | -19,05 |
| 11 | 21 | 16 | -23,81 |
| 12 | 33 | 14 | -57,58 |
| 13 | 24 | 14 | -41,67 |
| 14 | 35 | 20 | -42,86 |
| 15 | 54 | 34 | -37,04 |
| 16 | 55 | 32 | -41,82 |
| 17 | 65 | 17 | -73,85 |

## DISCUSSION

This study prospectively evaluated the impact of two different surgical approaches for treating VV associated with GSV insufficiency: conventional surgery (which involves saphenectomy with collateral VV resection) and isolated saphenectomy. The methodology used allowed for temporal control of clinical variables and systematic data collection at three different time points, making it possible to analyze the interventions effectively. This analysis demonstrated that isolated treatment of the GSV can lead to a comparable improvement in symptoms and quality of life, with an approximate 46% reduction in the volume of VV.

The CEAP classification assessment (as presented in Table 3), which evaluates the clinical, etiological, anatomical, and pathophysiological situation and was developed to standardize patients, shows that, before treatment, many patients were in moderate stages (C2, C3), including some in more advanced stages (C4, C6). After the interventions, there was a significant migration of patients to less severe stages, especially to stages C1 and C2, suggesting that the treatment has a clinically relevant effect. However, since the p-values between the groups did not show significant differences in either the pre- or postoperative periods, we did not observe superiority of one approach over the other in this specific assessment.

In terms of quality of life, preoperative scores indicated differences between the groups. The conventional group had higher (better) scores in the VEINES-QoL and VEINES-Sym domains, reflecting better baseline clinical status. This may be related to lower rates of active smoking and lower prevalence of hypertension in this cohort. Despite these differences, both groups showed significant improvement in quality of life scores after surgery (p < 0.005), with stable results up to the sixth month of follow-up. This demonstrates equivalent results at least in the short and medium terms. This observation is clinically relevant, especially in contexts where access to complete treatment is limited by resources.

The assessment of the volume of VV in the lower limbs showed that isolated GSV treatment resulted in a reduction of approximately 46% in dilated veins, which is a clinically relevant benefit. In patients with a lower VV load, abbreviated treatment eliminated up to 90% of the affected veins. As clinical evolution was similar across both groups, with equivalent CEAP reclassification 6 months post-treatment, it is suggested that collateral vein removal may not be necessary in some cases. This reinforces the hypothesis that interrupting GSV reflux may be sufficient to induce both functional and aesthetic regression of collateral veins. In our study, the regression ranged from 20% to 90%, indicating that the individual responses to treatment may depend not only on the number of preoperative veins, but also on their anatomical locations and the patient’s clinical conditions.

Existing literature supports the idea that thermoablative treatment of the GSV without simultaneous treatment of VV can benefit patients. Initial studies on this subject, dating back to the early 2000s, showed that exclusive radiofrequency treatment of the GSV promoted complete resolution of varicose veins in 13% of limbs, while unresolved VV reduced in diameter by an average of approximately 35%.^18^ In addition, exclusive treatment of the GSV was able to postpone or even eliminate the need for subsequent procedures.^19^

Recent studies, including randomized clinical trials, show a similar trend: combined treatment (saphenous vein + VV), while potentially offering better short-term benefit (which was not observed in our study), may lead to more complications (such as thromboembolic events) and, in the long term, show results comparable to abbreviated treatment^20^. Moreover, even if further treatment of the VV is necessary later, prior isolated saphenous vein ablation may result in smaller phlebectomy procedures, resulting in lower costs and fewer complications associated with a second procedure.^13^ Similar findings have also been observed with the use of mechanochemical ablation techniques: isolated saphenous vein treatment promotes a patient satisfaction rate equivalent to cases in which it was complemented with phlebectomy in the same procedure.^12^

Isolated ablation of the GSV has significant anatomical and hemodynamic effects on its tributaries. After treating saphenous reflux, the mean diameter of collateral veins is significantly reduced, and most of the initially insufficient tributaries return to normal after treatment of the truncal vein^21,22^. However, in EVTA, this effect could be secondary to the thermal component. Our study suggests that this is probably due to the mechanical effect of eliminating the reflux itself.

Although ablative techniques have become the gold standard in treating GSV insufficiency, access to this therapeutic modality remains limited in many parts of the world. In Latin America, for example, endovascular techniques remain inaccessible to a significant portion of the population.^16^ Even in developed countries in Europe and North America, there are still geographic and socioeconomic disparities in individuals’ access to less invasive treatment modalities^23–25^.

Furthermore, in low- and middle-income countries, patients with VV often experience delayed access to specialized care. These individuals usually receive treatment at an older age and present with more severe and numerous VV, often accompanied by complications such as venous ulcers.^26^ In this context, it is reasonable to seek alternatives that enable abbreviated conventional surgeries, which have a lower impact on patients and allow treatment for a larger number of individuals, all while maintaining the patient’s perception of quality of life.

Extensive literature demonstrates that conventional surgery is comparable to minimally invasive techniques in terms of technical success and in promoting equivalent and sustained improvement in quality of life. ^27–31^ In a context like Brazil’s, where the majority of the population depends on the public health system and often consults specialist at an advanced age with significant VV, the application of abbreviated procedures can enhance quality of life. Moreover, these procedures can guarantee lower perioperative morbidity and a reduced need for adjuvant procedures. This makes abbreviated surgeries an excellent alternative for both patients and the healthcare system, and they could be adopted as a health policy to ensure lower costs and faster treatment for a larger number of individuals.^32^

### Limitations

Although the present study was a prospective trial, it was conducted at a single center and did not involve a randomization process. While participant allocation was not randomized, the stratification followed institutional practices, ensuring that all patients received standard treatment by the end of the study. However, this approach may have introduced some selection bias, as patients with bilateral involvement may exhibit more severe clinical symptoms and, therefore, have different profiles compared to those with unilateral involvement.

The VEINES score, used to assess symptoms and quality of life, is typically read and completed by the patients themselves. However, given that this is a public service, where many of the patients have lower socioeconomic backgrounds, it was decided that the questionnaire would be administered by the physician, who would ask the patients all the questions, which may lead to biases associated with filling out the questionnaire.

The methodology used to quantify VV and compare different time points shows a reduction in the volume of visible veins in the abbreviated procedure. However, this assessment lacks objective measurement, such as ultrasound, which raises concerns about its analytical value.

Additionally, in both pre- and postoperative assessments, the evaluators were not blinded, which may have introduced biases into this assessment.

## CONCLUSION

Isolated saphenectomy and the conventional technique offer benefits for up to 6 months in terms of clinical and quality of life improvement in the treatments for patients suffering from VV of the lower limbs associated with GSV insufficiency.

The conventional technique enables the removal of the saphenous vein, along with 100% of the collateral VV. In comparison, isolated saphenectomy only removes the saphenous vein, leading to a 46% reduction in dilated veins.

These findings have significant implications for clinical practice, particularly in resource-limited settings, such as health systems in low- and middle-income countries, where access to minimally invasive techniques is limited and patients face greater challenges in accessing healthcare services. Adopting this abbreviated approach enables more patients to be treated with reduced perioperative morbidity. This may expand access to interventions for the treatment of VV, potentially reducing the incidence of complications, and generating cost savings for health systems.

## Data Availability

All data produced in the present study are available upon reasonable request to the authors

